# Prevalence, co-occurrence, and associated factors of post-dengue depressive, anxiety, and stress symptoms among adults in Bangladesh: a cross-sectional study

**DOI:** 10.64898/2026.11.26.26369032

**Authors:** Abdullah Al Mamun Sarker, Fahmida Sarker, Sadia Islam, Rowshan Ara Afrin, Atiqul Haq Mazumder, Zeenat Farzana Rahman, Sohrab Alam

**Author notes:** Corresponding author: Abdullah Al Mamun Sarker. These authors contributed equally and share first authorship (Abdullah Al Mamun Sarker, Fahmida Sarker). These authors contributed equally and share senior authorship (Atiqul Haq Mazumder, Zeenat Farzana Rahman, Sohrab Alam).

## Abstract

**Background:** Evidence on psychological symptoms following dengue remains limited, particularly in Bangladesh despite its recurrent dengue burden. We aimed to estimate the prevalence and co-occurrence of depressive, anxiety, and stress symptoms after dengue hospitalisation and to identify sociodemographic, behavioural, and clinical factors associated with these outcomes.

**Methods:** We conducted a cross-sectional study among adults aged 18–60 years with confirmed dengue admitted to BIRDEM General Hospital in Dhaka, Bangladesh, between June and October 2024. Patients with pre-existing hypertension, diabetes, cardiovascular disease, cancer, or respiratory disease were excluded. Eligible patients consented during hospitalisation and were contacted approximately one month after discharge for telephone assessment using the Bangla Depression Anxiety Stress Scales-21 (DASS-21). Moderate-or-greater symptoms were defined as scores ≥14 for depression, ≥10 for anxiety, and ≥19 for stress. Associations were estimated using multivariable logistic regression with 12 prespecified covariates.

**Results:** Of 397 patients approached, 338 agreed to screening, 306 were eligible and consented to follow-up, and 288 completed assessments (94.1% of eligible participants). Depressive, anxiety, and stress symptoms were present in 126 (43.75%), 129 (44.79%), and 91 (31.60%) participants, respectively; 72.57% had at least one outcome. Depression was associated with age 45–60 years (adjusted odds ratio [AOR] 2.28, 95% CI 1.12–4.69), graduate-level education or above (2.94, 1.52–5.80), and being married (1.79, 1.03–3.17). Anxiety was associated with age 30–44 years (2.00, 1.11–3.65), widowed/divorced status (4.17, 1.65–11.26), and previous mental-health illness (2.39, 1.18–4.97). Stress was associated with smoking (3.94, 2.09–7.61), middle income (3.11, 1.61–6.16), and rural residence (2.25, 1.24–4.13).

**Conclusions:** Depressive, anxiety, and stress symptoms were common approximately one month after dengue hospitalisation and were associated with distinct sociodemographic, behavioural, and clinical characteristics. These findings support greater attention to psychological well-being during post-dengue care.

## Introduction

Dengue is an expanding mosquito-borne neglected tropical disease [1]. Recent estimates reported approximately 14.1 million cases and 9,508 deaths worldwide in 2024 [2]. Over three decades, dengue-related disability-adjusted life-years (DALYs) increased from 1.25 million to 2.08 million [3]. Bangladesh has experienced recurrent outbreaks, including a record 321 179 cases and 1705 deaths in 2023 [4,5]. Beyond its acute clinical symptoms, dengue has been associated with psychological and psychiatric morbidity during and after infection. Reviews of the available evidence have documented depression, anxiety, stress, sleep disturbances, and other psychiatric manifestations across the acute and post-acute phases of dengue [6–8]. Evidence from eight observational studies identified occurrence of depression and anxiety during both acute and convalescent stages [6], while subsequent reviews have described a broader range of psychiatric and psychological symptoms [7,8]. A 2025 scoping review identified 14 studies examining psychological outcomes of dengue, demonstrating both short-term and longer-term psychological morbidity [8]. Evidence from South Asia also suggests psychological symptoms following dengue infection. In Pakistan, anxiety and depressive symptoms were reported among patients with dengue [9]. In Sri Lanka, adults assessed 6–24 months after dengue had significantly higher depression, anxiety, and stress scores than age-sex-matched controls [10]. Longitudinal evidence further suggests that psychological symptoms extend beyond acute illness after dengue. A nationwide population-based cohort study from Taiwan found an increased risk of depressive disorders within 3 months after dengue. They found a 90% higher subdistribution hazard among the hospitalised dengue patients [11]. A prospective cohort study in Ecuador reported that 44.4% of adults with previous dengue reported a 2.7-fold higher risk of depressive symptoms and 87% higher risk of stress [12]. Despite Bangladesh’s recurrent dengue burden, evidence on psychological symptoms following dengue hospitalisation remains limited, particularly regarding the co-occurrence of these symptoms and the sociodemographic, behavioural, and clinical factors associated with them. Therefore, this study aimed to determine the prevalence and co-occurrence of depressive, anxiety, and stress symptoms among adults following hospitalisation for dengue in Bangladesh and to identify sociodemographic, behavioural, and clinical factors associated with these psychological outcomes.

## Materials and methods

### Study design and setting

A cross-sectional study was conducted among patients hospitalised with confirmed dengue infection at the Bangladesh Institute of Research and Rehabilitation in Diabetes, Endocrine and Metabolic Disorders (BIRDEM) General Hospital, Dhaka, Bangladesh, between June and October 2024. No formal a priori sample-size calculation was performed. Participants were recruited using consecutive sampling during the study period. Eligibility criteria included being aged 18–60 years, having confirmed dengue infection (based on a positive non-structural protein 1 [NS1] antigen test) requiring hospitalisation, and agreeing to participate in the planned post-discharge assessment. Patients with pre-existing chronic physical diseases, including hypertension, cancer, respiratory diseases, diabetes, and cardiovascular disease, were excluded.

Of 397 patients approached, 338 agreed to screening (85.1%); 32 were excluded (13 outside the eligible age range and 19 with pre-existing chronic physical diseases). The remaining 306 consented to telephone follow-up approximately one month after discharge; 18 were unreachable despite repeated attempts, leaving 288 participants (Fig 1). Sociodemographic, behavioural, clinical, and DASS-21 data were collected by telephone by trained psychological research personnel from Thikana Psychiatric and Drug Addiction Clinic, Mohammadpur, Dhaka. Psychologists from the same clinic reviewed completed assessments for completeness and scoring accuracy.

**Fig 1.**
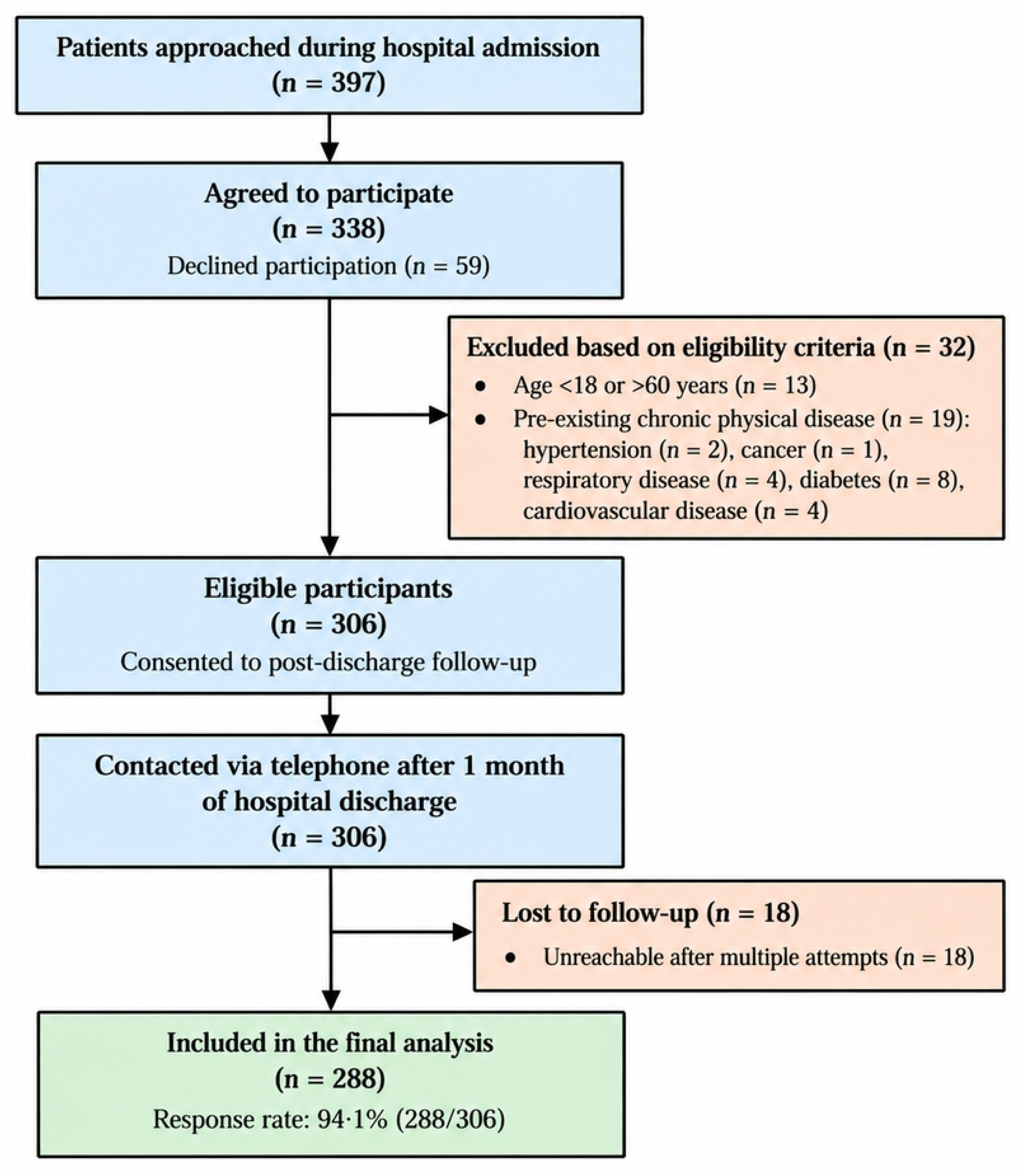
Flow of participants from initial approach to post-discharge telephone assessment.

### Study variables

The dependent variables were depression, anxiety, and stress symptoms, assessed approximately one month after discharge using the validated Bangla version of the DASS-21 [13]. The DASS-21 measures psychological symptoms rather than providing clinical psychiatric diagnoses and comprises three seven-item subscales for depression, anxiety, and stress. Each item was rated from 0 (did not apply to me at all) to 3 (applied to me very much, or most of the time) according to participants’ experiences during the preceding week. Items within each subscale were summed and multiplied by two. Depression scores were classified as normal (0–9), mild (10–13), moderate (14–20), severe (21–27), or extremely severe (≥28); anxiety as normal (0–7), mild (8–9), moderate (10–14), severe (15–19), or extremely severe (≥20); and stress as normal (0–14), mild (15–18), moderate (19–25), severe (26–33), or extremely severe (≥34). The corresponding thresholds for moderate-or-greater symptoms were ≥14 for depression, ≥10 for anxiety, and ≥19 for stress.

Independent variables included age group (18–29, 30–44, or 45–60 years), sex (female or male), educational attainment (primary or below, secondary, or graduate or above), monthly income (low (<BDT 20,000), middle (BDT 20,000–40,000), or high (>BDT 40,000)) [14], marital status (unmarried, married, or widowed/divorced), occupation (employed, unemployed/homemaker, student, or other), and residence (urban or rural). Behavioural and clinical variables included self-reported current smoking status (yes or no), self-reported history of COVID-19 infection and mental-health illness before the current dengue infection (yes or no), duration of hospital stay, and month of dengue infection (June–October). Hospital stay was categorised as short (1–3 days), moderate (4–6 days), or long (≥7 days).

### Statistical analysis

Characteristics were summarised as frequencies and percentages; DASS-21 subscale scores as means and SDs. Moderate-or-greater symptoms were compared with normal/mild symptoms for each binary outcome in regression and co-occurrence analyses. Symptom overlap was visualised using a Venn diagram. Unadjusted differences in psychological outcomes across sociodemographic, behavioural, and clinical characteristics were examined using Pearson’s χ² test. Separate univariable logistic regression models were fitted for each outcome to estimate crude associations with individual covariates, reported as crude odds ratios (CORs) with 95% CIs and p values. Separate multivariable logistic regression models were then fitted for depression, anxiety, and stress. All 12 prespecified covariates including age group, sex, education, monthly income, marital status, occupation, residence, smoking, hospital-stay category, month of dengue infection, previous COVID-19 infection, and previous mental-health illness were entered simultaneously into each model irrespective of statistical significance in the unadjusted analyses. Adjusted associations were reported as adjusted odds ratios (AORs) with 95% CIs and p values. There were no missing data for the variables included in the analyses among the 288 participants who completed follow-up; therefore, no imputation was required. DASS-21 internal consistency was assessed using Cronbach’s alpha. Multicollinearity was evaluated using generalised variance inflation factor (GVIF) diagnostics, and model calibration using the Hosmer–Lemeshow goodness-of-fit test. All tests were two-sided, with p < 0.05 considered statistically significant. Analyses were conducted using R version 4.3.1.

### Ethics statement

Ethical approval was obtained from the Ethical Review Committee (ERC) of the Diabetic Association of Bangladesh (BADAS) (BADAS-ERC/EC/24/08). Participants were informed about the study and planned post-discharge follow-up, and written informed consent was obtained before participation. Participation was voluntary, and all collected information was kept confidential and used solely for research purposes.

## Results

Of 306 eligible participants, 288 completed follow-ups (94.1%); 18 were unreachable. The median interval between hospital discharge and follow-up assessment was 32 days (IQR 30–33 days; range 30–35 days). Participants were predominantly aged 30–44 years (44.79%), female (56.60%), and married (52.43%). Low income was reported by 43.75%, and urban and rural participants were equally represented. Short hospital stays accounted for 46.18%; infections were most frequent in August (28.47%) and September (26.04%). Previous COVID-19 infection and mental-health illness were reported by 32.29% and 15.63%, respectively (Table 1).

**Table 1.** Sociodemographic, behavioural, and clinical characteristics of participants (N = 288).

| **Characteristic** | **n (%)** |
|---|---|
| **Age group, years** |  |
| 18–29 | 103 (35.76%) |
| 30–44 | 129 (44.79%) |
| 45–60 | 56 (19.44%) |
| **Sex** |  |
| Female | 163 (56.60%) |
| Male | 125 (43.40%) |
| **Educational attainment** |  |
| Primary or below | 100 (34.72%) |
| Secondary | 110 (38.19%) |
| Graduate or above | 78 (27.08%) |
| **Monthly income** |  |
| Low | 126 (43.75%) |
| Middle | 99 (34.38%) |
| High | 63 (21.88%) |
| **Marital status** |  |
| Unmarried | 106 (36.81%) |
| Married | 151 (52.43%) |
| Widowed/divorced | 31 (10.76%) |
| **Occupation** |  |
| Employed | 99 (34.38%) |
| Unemployed/homemaker | 97 (33.68%) |
| **Age group, years** |  |
| Student | 58 (20.14%) |
| Other | 34 (11.81%) |
| **Place of residence** |  |
| Urban | 144 (50.00%) |
| Rural | 144 (50.00%) |
| **Smoking status** |  |
| Yes | 75 (26.04%) |
| No | 213 (73.96%) |
| **Duration of hospital stay** |  |
| Short (1–3 days) | 133 (46.18%) |
| Moderate (4–6 days) | 103 (35.76%) |
| Long (≥7 days) | 52 (18.06%) |
| **Month of dengue infection** |  |
| June | 43 (14.93%) |
| July | 61 (21.18%) |
| August | 82 (28.47%) |
| September | 75 (26.04%) |
| October | 27 (9.38%) |
| **Previous COVID-19 infection** |  |
| Yes | 93 (32.29%) |
| No | 195 (67.71%) |
| **Previous mental-health illness** |  |
| Yes | 45 (15.63%) |
| No | 243 (84.38%) |

Values are n (%). Monthly income was categorized as low (<BDT 20,000), middle (BDT 20,000–40,000), and high (>BDT 40,000).

### Prevalence and co-occurrence of depressive, anxiety, and stress symptoms

The mean DASS-21 depression, anxiety, and stress scores were 11.78 (standard deviation [SD] 9.13), 10.86 (SD 9.42), and 10.78 (SD 8.85), respectively. Based on the DASS-21 cut-off scores, depressive symptoms were identified in 43.75% of participants, anxiety symptoms in 44.79%, and stress symptoms in 31.60%. Overall, 72.57% experienced at least one of the three symptoms. Fig 2 presents their co-occurrence. Depressive and anxiety symptoms without stress were reported by 12.2% of participants, depressive and stress symptoms without anxiety by 6.9%, and anxiety and stress symptoms without depression by 5.6%. All three symptoms occurred concurrently in 11.5% of participants, while 27.4% experienced none of the three symptoms.

**Fig 2.**
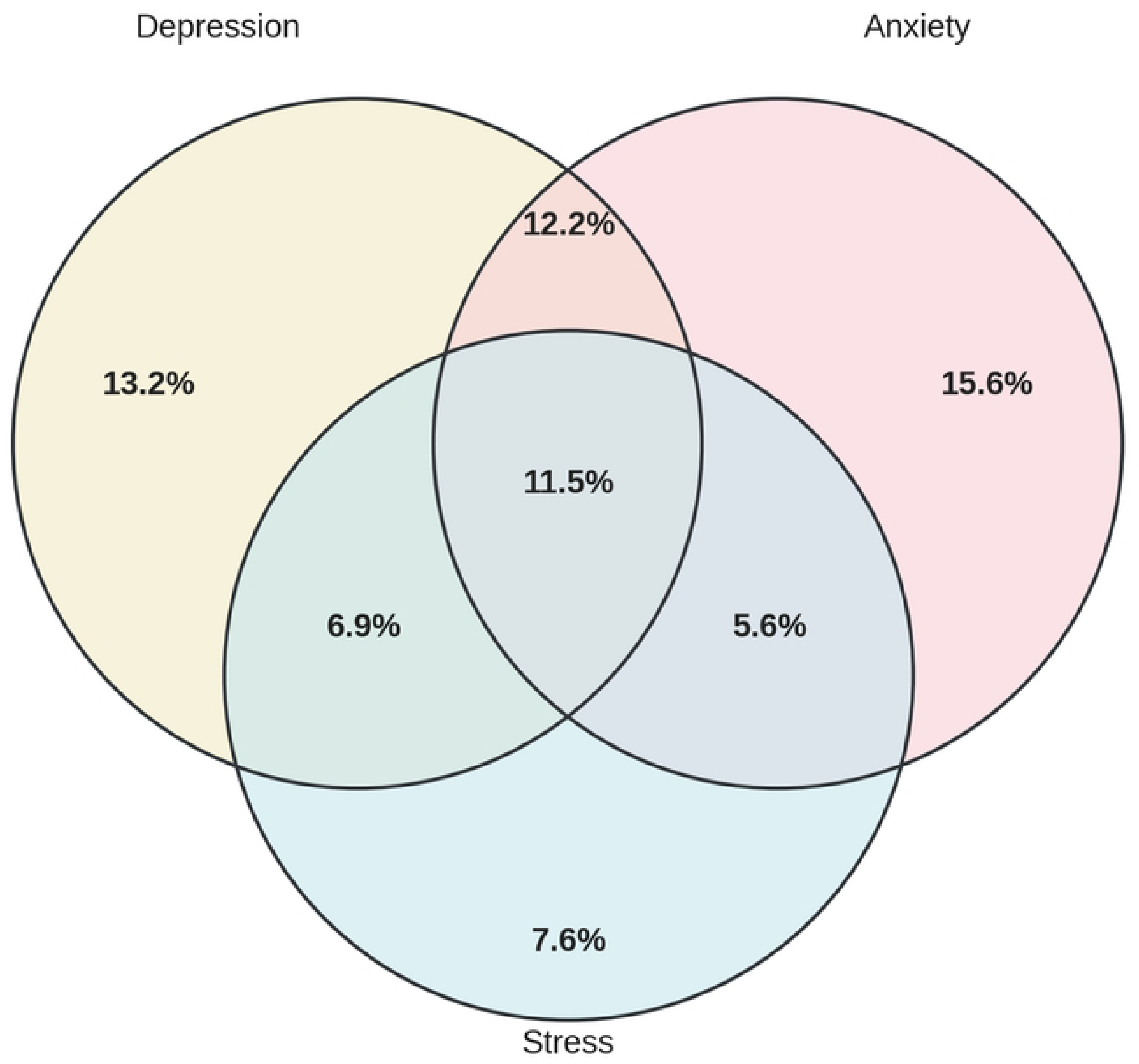
Co-occurrence of moderate-or-greater depressive, anxiety, and stress symptoms among 288 adults.

In the univariate comparisons, educational attainment and marital status differed significantly according to depressive symptom status (p = 0.003 and p = 0.023, respectively). For anxiety, significant differences were observed by sex (p = 0.012), marital status (p = 0.023), and month of dengue infection (p = 0.041). Stress status differed significantly by monthly income (p = 0.019), place of residence (p = 0.023), smoking status (p < 0.001), hospital-stay category (p = 0.019), and previous COVID-19 infection (p = 0.003). No statistically significant differences were observed for the remaining characteristics across the respective outcomes (Table 2).

**Table 2.** Univariate comparisons of participant characteristics by depressive, anxiety, and stress symptom status.

| Characteristic | Depression<br>Depression |  | Anxiety<br>Anxiety |  | Stress<br>Stress |  |
|---|---|---|---|---|---|---|
|  | No / Yes, n (%) | p value | No / Yes, n (%) | p value | No / Yes, n (%) | p value |
| **Age group** |  | **0.080** |  | **0.14** |  | **0.79** |
| 18–29 | 65 (40.12%) / 38 (30.16%) |  | 61 (38.36%) / 42 (32.56%) |  | 73 (37.06%) / 30 (32.97%) |  |
| 30–44 | 72 (44.44%) / 57 (45.24%) |  | 63 (39.62%) / 66 (51.16%) |  | 86 (43.65%) / 43 (47.25%) |  |
| 45–60 | 25 (15.43%) / 31 (24.60%) |  | 35 (22.01%) / 21 (16.28%) |  | 38 (19.29%) / 18 (19.78%) |  |
| **Sex** |  | **0.78** |  | **0.012** |  | **0.80** |
| Female | 90 (55.56%) / 73 (57.94%) |  | 79 (49.69%) / 84 (65.12%) |  | 110 (55.84%) / 53 (58.24%) |  |
| Male | 72 (44.44%) / 53 (42.06%) |  | 80 (50.31%) / 45 (34.88%) |  | 87 (44.16%) / 38 (41.76%) |  |
| **Educational attainment** |  | **0.003** |  | **0.20** |  | **0.79** |
| Primary or below | 63 (38.89%) / 37 (29.37%) |  | 48 (30.19%) / 52 (40.31%) |  | 70 (35.53%) / 30 (32.97%) |  |
| Secondary | 68 (41.98%) / 42 (33.33%) |  | 65 (40.88%) / 45 (34.88%) |  | 76 (38.58%) / 34 (37.36%) |  |
| Graduate or above | 31 (19.14%) / 47 (37.30%) |  | 46 (28.93%) / 32 (24.81%) |  | 51 (25.89%) / 27 (29.67%) |  |
| **Monthly income** |  | **0.46** |  | **0.91** |  | **0.019** |
| Low | 76 (46.91%) / 50 (39.68%) |  | 68 (42.77%) / 58 (44.96%) |  | 96 (48.73%) / 30 (32.97%) |  |
| Middle | 52 (32.10%) / 47 (37.30%) |  | 55 (34.59%) / 44 (34.11%) |  | 58 (29.44%) / 41 (45.05%) |  |
| High | 34 (20.99%) / 29 (23.02%) |  | 36 (22.64%) / 27 (20.93%) |  | 43 (21.83%) / 20 (21.98%) |  |
| **Marital status** |  | **0.023** |  | **0.023** |  | **0.66** |
| Unmarried | 66 (40.74%) / 40 (31.75%) |  | 63 (39.62%) / 43 (33.33%) |  | 73 (37.06%) / 33 (36.26%) |  |
| Married | 74 (45.68%) / 77 (61.11%) |  | 86 (54.09%) / 65 (50.39%) |  | 105 (53.30%) / 46 (50.55%) |  |
| Widowed/divorced | 22 (13.58%) / 9 (7.14%) |  | 10 (6.29%) / 21 (16.28%) |  | 19 (9.64%) / 12 (13.19%) |  |
| **Occupation** |  | **0.64** |  | **0.20** |  | **0.37** |
| Employed | 55 (33.95%) / 44 (34.92%) |  | 62 (38.99%) / 37 (28.68%) |  | 68 (34.52%) / 31 (34.07%) |  |
| Unemployed/homemaker | 58 (35.80%) / 39 (30.95%) |  | 46 (28.93%) / 51 (39.53%) |  | 70 (35.53%) / 27 (29.67%) |  |
| Student | 33 (20.37%) / 25 (19.84%) |  | 32 (20.13%) / 26 (20.16%) |  | 40 (20.30%) / 18 (19.78%) |  |
| Other | 16 (9.88%) / 18 (14.29%) |  | 19 (11.95%) / 15 (11.63%) |  | 19 (9.64%) / 15 (16.48%) |  |
| **Place of residence** |  | **0.55** |  | **0.81** |  | **0.023** |
| Urban | 78 (48.15%) / 66 (52.38%) |  | 81 (50.94%) / 63 (48.84%) |  | 108 (54.82%) / 36 (39.56%) |  |
| Rural | 84 (51.85%) / 60 (47.62%) |  | 78 (49.06%) / 66 (51.16%) |  | 89 (45.18%) / 55 (60.44%) |  |
| **Smoking status** |  | **0.070** |  | **0.27** |  | **<0.001** |
| Yes | 35 (21.60%) / 40 (31.75%) |  | 46 (28.93%) / 29 (22.48%) |  | 38 (19.29%) / 37 (40.66%) |  |
| **Hospital-stay category** |  | **0.69** |  | **0.48** |  | **0.019** |
| Short stay | 78 (48.15%) / 55 (43.65%) |  | 74 (46.54%) / 59 (45.74%) |  | 102 (51.78%) / 31 (34.07%) |  |
| Moderate stay | 57 (35.19%) / 46 (36.51%) |  | 60 (37.74%) / 43 (33.33%) |  | 64 (32.49%) / 39 (42.86%) |  |
| Long stay | 27 (16.67%) / 25 (19.84%) |  | 25 (15.72%) / 27 (20.93%) |  | 31 (15.74%) / 21 (23.08%) |  |
| **Month of infection** |  | **0.31** |  | **0.041** |  | **0.78** |
| June | 26 (16.05%) / 17 (13.49%) |  | 27 (16.98%) / 16 (12.40%) |  | 32 (16.24%) / 11 (12.09%) |  |
| July | 29 (17.90%) / 32 (25.40%) |  | 28 (17.61%) / 33 (25.58%) |  | 39 (19.80%) / 22 (24.18%) |  |
| August | 48 (29.63%) / 34 (26.98%) |  | 38 (23.90%) / 44 (34.11%) |  | 54 (27.41%) / 28 (30.77%) |  |
| September | 40 (24.69%) / 35 (27.78%) |  | 47 (29.56%) / 28 (21.71%) |  | 53 (26.90%) / 22 (24.18%) |  |
| October | 19 (11.73%) / 8 (6.35%) |  | 19 (11.95%) / 8 (6.20%) |  | 19 (9.64%) / 8 (8.79%) |  |
| **Previous COVID-19 infection** |  | **0.47** |  | **0.83** |  | **0.003** |
| Yes | 49 (30.25%) / 44 (34.92%) |  | 50 (31.45%) / 43 (33.33%) |  | 52 (26.40%) / 41 (45.05%) |  |
| **Previous mental-health illness** |  | **0.057** |  | **0.081** |  | **0.55** |
| Yes | 19 (11.73%) / 26 (20.63%) |  | 19 (11.95%) / 26 (20.16%) |  | 33 (16.75%) / 12 (13.19%) |  |

Data are n (%). Within each outcome, values are shown as No / Yes. p values are from Pearson's chi-square tests. No and Yes refer to absence or presence of the respective moderate-or-greater DASS-21 outcome.

In univariable logistic regression analyses, participants aged 45–60 years had higher odds of depressive symptoms than those aged 18–29 years (COR 2.12, 95% CI 1.10–4.14; p = 0.026). Graduate-level education or above was associated with higher odds of depressive symptoms compared with primary education or below (COR 2.58, 95% CI 1.41–4.78; p = 0.002), as were being married (COR 1.72, 95% CI 1.04–2.86; p = 0.036) and having a previous mental-health illness (COR 1.96, 95% CI 1.03–3.77; p = 0.041). For anxiety, males had lower odds than females (COR 0.53, 95% CI 0.33–0.85; p = 0.009), whereas widowed/divorced participants (COR 3.08, 95% CI 1.35–7.43; p = 0.009) and unemployed participants or homemakers (COR 1.86, 95% CI 1.05–3.30; p = 0.033) had higher odds. Higher odds of stress symptoms were observed among participants with middle income (COR 2.26, 95% CI 1.28–4.04; p = 0.005), rural residence (COR 1.85, 95% CI 1.12–3.09; p = 0.017), smoking (COR 2.87, 95% CI 1.66–4.98; p < 0.001), moderate (COR 2.01, 95% CI 1.14–3.55; p = 0.016) or long hospital stays (COR 2.23, 95% CI 1.12–4.42; p = 0.022), and previous COVID-19 infection (COR 2.29, 95% CI 1.36–3.86; p = 0.002) (Table 3).

**Table 3.** Univariable logistic regression analyses of factors associated with depressive, anxiety, and stress symptoms.

| **Variable / category<br>Variable / category** |  | **Depression<br>Depression** |  | **Anxiety<br>Anxiety** |  | **Stress<br>Stress** |  |
|  |  | **COR (95% CI)** | **p value** | **COR (95% CI)** | **p value** | **COR (95% CI)** | **p value** |
|---|---|---|---|---|---|---|---|
| Age group | 30–44 | 1.35 (0.80–2.31) | 0.262 | 1.52 (0.90–2.58) | 0.116 | 1.22 (0.70–2.14) | 0.493 |
|  | 45–60 | 2.12 (1.10–4.14) | 0.026 | 0.87 (0.44–1.69) | 0.687 | 1.15 (0.56–2.32) | 0.692 |
| Sex | Male | 0.91 (0.57–1.45) | 0.686 | 0.53 (0.33–0.85) | 0.009 | 0.91 (0.55–1.50) | 0.702 |
| Education | Secondary | 1.05 (0.60–1.84) | 0.860 | 0.64 (0.37–1.10) | 0.108 | 1.04 (0.58–1.89) | 0.886 |
|  | Graduate or above | 2.58 (1.41–4.78) | 0.002 | 0.64 (0.35–1.16) | 0.146 | 1.24 (0.65–2.33) | 0.513 |
| Monthly income | Middle | 1.37 (0.81–2.34) | 0.242 | 0.94 (0.55–1.59) | 0.812 | 2.26 (1.28–4.04) | 0.005 |
|  | High | 1.30 (0.70–2.39) | 0.405 | 0.88 (0.48–1.61) | 0.679 | 1.49 (0.76–2.90) | 0.245 |
| Marital status | Married | 1.72 (1.04–2.86) | 0.036 | 1.11 (0.67–1.84) | 0.692 | 0.97 (0.57–1.67) | 0.909 |
|  | Widowed/divorced | 0.68 (0.27–1.57) | 0.376 | 3.08 (1.35–7.43) | 0.009 | 1.40 (0.60–3.19) | 0.431 |
| Occupation | Unemployed/homemaker | 0.84 (0.48–1.48) | 0.548 | 1.86 (1.05–3.30) | 0.033 | 0.85 (0.46–1.56) | 0.594 |
|  | Student | 0.95 (0.49–1.82) | 0.870 | 1.36 (0.70–2.63) | 0.358 | 0.99 (0.48–1.98) | 0.971 |
|  | Other | 1.41 (0.64–3.10) | 0.393 | 1.32 (0.59–2.91) | 0.488 | 1.73 (0.77–3.85) | 0.178 |
| Place of residence | Rural | 0.84 (0.53–1.34) | 0.476 | 1.09 (0.68–1.73) | 0.722 | 1.85 (1.12–3.09) | 0.017 |
| Smoking | Yes | 1.69 (0.99–2.88) | 0.053 | 0.71 (0.41–1.21) | 0.216 | 2.87 (1.66–4.98) | <0.001 |
| Hospital stay | Moderate stay | 1.14 (0.68–1.93) | 0.611 | 0.90 (0.53–1.51) | 0.688 | 2.01 (1.14–3.55) | 0.016 |
|  | Long stay | 1.31 (0.69–2.51) | 0.407 | 1.35 (0.71–2.59) | 0.355 | 2.23 (1.12–4.42) | 0.022 |
| Month of infection | July | 1.69 (0.77–3.77) | 0.195 | 1.99 (0.90–4.48) | 0.091 | 1.64 (0.70–3.99) | 0.260 |
|  | August | 1.08 (0.51–2.32) | 0.835 | 1.95 (0.93–4.22) | 0.082 | 1.51 (0.67–3.54) | 0.328 |
|  | September | 1.34 (0.63–2.90) | 0.453 | 1.01 (0.46–2.21) | 0.989 | 1.21 (0.53–2.89) | 0.662 |
|  | October | 0.64 (0.22–1.77) | 0.401 | 0.71 (0.24–1.96) | 0.516 | 1.22 (0.41–3.58) | 0.711 |
| Previous COVID-19 infection | Yes | 1.24 (0.75–2.03) | 0.400 | 1.09 (0.66–1.79) | 0.733 | 2.29 (1.36–3.86) | 0.002 |
| Previous mental-health illness | Yes | 1.96 (1.03–3.77) | 0.041 | 1.86 (0.98–3.58) | 0.059 | 0.75 (0.36–1.50) | 0.440 |

COR=crude odds ratio; CI=confidence interval. Reference categories: age 18–29 years; female; primary education or below; low income; unmarried; employed; urban residence; non-smoker; short hospital stay; June infection; no previous COVID-19 infection; no previous mental-health illness. p<0.05 was considered statistically significant.

After multivariable adjustment, participants aged 45–60 years had higher odds of depressive symptoms than those aged 18–29 years (AOR 2.28, 95% CI 1.12–4.69; p = 0.023). Graduate-level education or above remained associated with higher odds of depressive symptoms (AOR 2.94, 95% CI 1.52–5.80; p = 0.002), as did being married (AOR 1.79, 95% CI 1.03–3.17; p = 0.042). For anxiety, participants aged 30–44 years had higher odds than those aged 18–29 years (AOR 2.00, 95% CI 1.11–3.65; p = 0.022), while males had lower odds than females (AOR 0.52, 95% CI 0.31–0.89; p = 0.018). Higher odds of anxiety symptoms were also observed among widowed/divorced participants (AOR 4.17, 95% CI 1.65–11.26; p = 0.003), unemployed participants or homemakers (AOR 1.91, 95% CI 1.02–3.61; p = 0.045), participants infected during August compared with June (AOR 2.45, 95% CI 1.06–5.82; p = 0.038), and those with a previous mental-health illness (AOR 2.39, 95% CI 1.18–4.97; p = 0.017). For stress, higher odds were observed among participants with middle income (AOR 3.11, 95% CI 1.61–6.16; p < 0.001), rural residence (AOR 2.25, 95% CI 1.24–4.13; p = 0.008), smoking (AOR 3.94, 95% CI 2.09–7.61; p < 0.001), moderate (AOR 2.71, 95% CI 1.40–5.34; p = 0.003) or long hospital stays (AOR 2.83, 95% CI 1.28–6.37; p = 0.011), and previous COVID-19 infection (AOR 3.29, 95% CI 1.78–6.22; p < 0.001) (Table 4).

**Table 4.** Multivariable logistic regression analyses of factors associated with depressive, anxiety, and stress symptoms.

| **Variable / category**<br>**Variable / category** |  | **Depression**<br>**Depression** |  | **Anxiety**<br>**Anxiety** |  | **Stress**<br>**Stress** |  |
|---|---|---|---|---|---|---|---|
|  |  | **AOR (95% CI)** | **p value** | **AOR (95% CI)** | **p value** | **AOR (95% CI)** | **p value** |
| Age group | 30–44 | 1.51 (0.84–2.75) | 0.168 | 2.00 (1.11–3.65) | 0.022 | 1.28 (0.67–2.46) | 0.456 |
|  | 45–60 | 2.28 (1.12–4.69) | 0.023 | 0.81 (0.38–1.71) | 0.584 | 1.32 (0.58–3.00) | 0.501 |
| Sex | Male | 0.90 (0.54–1.53) | 0.707 | 0.52 (0.31–0.89) | 0.018 | 0.76 (0.42–1.35) | 0.355 |
| Educational attainment | Secondary | 1.21 (0.66–2.22) | 0.543 | 0.67 (0.37–1.23) | 0.199 | 0.91 (0.46–1.82) | 0.797 |
|  | Graduate or above | 2.94 (1.52–5.80) | 0.002 | 0.63 (0.32–1.22) | 0.169 | 1.12 (0.54–2.33) | 0.762 |
| Monthly income | Middle | 1.74 (0.96–3.17) | 0.069 | 0.91 (0.50–1.65) | 0.756 | 3.11 (1.61–6.16) | <0.001 |
|  | High | 1.35 (0.69–2.67) | 0.380 | 0.95 (0.48–1.89) | 0.894 | 1.67 (0.76–3.65) | 0.201 |
| Marital status | Married | 1.79 (1.03–3.17) | 0.042 | 1.27 (0.72–2.25) | 0.407 | 1.02 (0.55–1.92) | 0.946 |
|  | Widowed/divorced | 0.77 (0.29–1.95) | 0.592 | 4.17 (1.65–11.26) | 0.003 | 1.82 (0.67–4.81) | 0.232 |
| Occupation | Unemployed/homemaker | 0.79 (0.42–1.48) | 0.469 | 1.91 (1.02–3.61) | 0.045 | 0.74 (0.36–1.51) | 0.411 |
|  | Student | 0.78 (0.37–1.62) | 0.515 | 1.11 (0.53–2.32) | 0.773 | 0.98 (0.43–2.19) | 0.960 |
|  | Other | 1.41 (0.59–3.40) | 0.434 | 1.10 (0.44–2.73) | 0.832 | 2.20 (0.84–5.75) | 0.105 |
| Place of residence | Rural | 0.88 (0.52–1.48) | 0.619 | 1.10 (0.65–1.86) | 0.731 | 2.25 (1.24–4.13) | 0.008 |
| Smoking | Yes | 1.64 (0.91–2.96) | 0.098 | 0.59 (0.32–1.08) | 0.090 | 3.94 (2.09–7.61) | <0.001 |
| Hospital stay | Moderate stay | 1.08 (0.60–1.92) | 0.807 | 1.03 (0.57–1.86) | 0.915 | 2.71 (1.40–5.34) | 0.003 |
|  | Long stay | 1.12 (0.55–2.27) | 0.762 | 1.48 (0.72–3.06) | 0.285 | 2.83 (1.28–6.37) | 0.011 |
| Month of infection | July | 1.69 (0.70–4.18) | 0.247 | 2.35 (0.96–5.93) | 0.065 | 1.39 (0.50–3.96) | 0.533 |
|  | August | 1.40 (0.61–3.29) | 0.434 | 2.45 (1.06–5.82) | 0.038 | 1.44 (0.56–3.86) | 0.456 |
|  | September | 1.39 (0.60–3.31) | 0.447 | 1.00 (0.42–2.41) | 0.995 | 1.52 (0.57–4.28) | 0.413 |
|  | October | 0.82 (0.26–2.44) | 0.718 | 0.67 (0.21–2.04) | 0.483 | 1.26 (0.35–4.43) | 0.718 |
| Previous COVID-19 infection | Yes | 1.32 (0.76–2.29) | 0.326 | 1.03 (0.59–1.80) | 0.914 | 3.29 (1.78–6.22) | <0.001 |
| Previous mental-health illness | Yes | 1.94 (0.97–3.92) | 0.062 | 2.39 (1.18–4.97) | 0.017 | 0.70 (0.30–1.53) | 0.378 |

AOR=adjusted odds ratio; CI=confidence interval. All 12 prespecified covariates were entered simultaneously into each model. Reference categories: age 18–29 years; female; primary education or below; low income; unmarried; employed; urban residence; non-smoker; short hospital stay; June infection; no previous COVID-19 infection; no previous mental-health illness. p<0.05 was considered statistically significant.

The DASS-21 demonstrated acceptable internal consistency, with Cronbach’s alpha coefficients of 0.767 for depression, 0.821 for anxiety, 0.771 for stress, and 0.784 for the overall DASS-21 scale. There was no evidence of substantial multicollinearity among the covariates, with all adjusted generalized variance inflation factor values ≤1.08 across the depression, anxiety, and stress regression models. The Hosmer– Lemeshow goodness-of-fit tests indicated adequate model calibration for depression (χ²=9.327, df=8, p = 0.316), anxiety (χ²=11.689, df=8, p = 0.166), and stress (χ²=3.628, df=8, p = 0.889).

## Discussion

We assessed the burden and associated factors of depressive, anxiety, and stress symptoms among dengue-positive patients in Bangladesh approximately one month after hospital discharge. Depression, anxiety, and stress symptoms were reported by 43.75%, 44.79%, and 31.60% of participants, respectively. Nearly three-quarters (72.57%) experienced at least one of these outcomes, and 11.5% experienced all three concurrently. After adjustment for the sociodemographic, behavioural, and clinical covariates included in the multivariable models, depressive symptoms were more common among older participants, those with graduate-level education or above, and married participants. Anxiety was associated with middle age, being widowed or divorced, unemployment or homemaking, infection during August, and previous mental-health illness, whereas men had lower odds than women. The strongest association was observed among widowed or divorced participants, who had more than four-times higher odds of anxiety (AOR 4.17, 95% CI 1.65–11.26). Stress was associated with middle income, rural residence, smoking, longer hospital stay, and previous COVID-19 infection; smoking showed the strongest association, with almost four-times higher odds of stress (AOR 3.94, 95% CI 2.09–7.61).

The global burden of dengue has increased substantially, with estimated incidence rising from 26.45 million cases in 1990 to 58.96 million in 2021, deaths from 14 315 to 29 075, and DALYs from 1.25 million to 2.08 million over the same period [2]. Although the acute clinical burden of dengue is well recognised [1], psychological outcomes during recovery have received considerably less attention. Existing studies of post-dengue mental health remain relatively limited and heterogeneous in their timing, outcome definitions, and assessment methods. Much of the available evidence has focused either on psychological symptoms during the acute phase or on longer-term psychiatric outcomes, whereas fewer studies have examined psychological morbidity during early recovery or shortly after hospital discharge [6–8,10–12].

This gap is particularly important because depression, anxiety, stress, fatigue, and other post-infectious symptoms have been reported during dengue recovery [7,8,12]. Recent population-based studies from Singapore have also identified increased risks of post-acute neuropsychiatric complications and associated longer-term disability following dengue infection, further highlighting the importance of investigating health outcomes beyond the acute illness [15,16].

Our prevalence estimates are consistent with previous studies, although differences in study design, timing, and outcome assessment limit direct comparison. In Ecuador, 44.4% of adults with dengue had elevated depressive symptoms three months after infection compared with 17.1% of matched controls, and stress was also more frequent among dengue cases [12]. A Sri Lankan case-control study similarly reported higher depression, anxiety, and stress among adults with previous dengue than among matched controls 6– 24 months after infection [10]. A large Taiwanese cohort also found increased risks of depressive disorders after dengue and short-term anxiety among hospitalised patients [11]. More recently, a hospital-based study from Bangladesh reported depressive symptoms in approximately 35% of 246 adults assessed within 1–3 months after discharge following dengue infection, providing additional evidence of psychological symptoms during the post-dengue period in a comparable setting [17].

Higher psychological symptom prevalence has also been reported during acute dengue. In Pakistan, approximately 60% of hospitalised patients had anxiety symptoms and 62.2% had depressive symptoms, with greater physical symptom severity associated with higher scores [9]. Another hospital-based Pakistani study reported substantial depression, anxiety, and stress [18]. More recent evidence from Sri Lanka identified stress approximately one month after dengue, with female sex, persistent headache, and myalgia associated with greater risk [19]. A 2026 systematic review further highlighted post-dengue fatigue and depression as understudied outcomes [20]. These findings indicate that psychological symptoms occur during both acute illness and recovery, although their frequency and associated factors vary across settings and follow-up periods.

Several factors could contribute to psychological symptoms after dengue. Acute illness is often accompanied by pain, weakness, sleep disturbance, and disruption of daily activities. A recent multicentre study among hospitalised dengue patients in Bangladesh documented considerable impairments in mobility, usual activities, pain or discomfort, and anxiety or depression, highlighting the broader physical and psychological burden of dengue during hospitalisation [21]. Greater symptom severity has been associated with higher depression and anxiety scores [9], while persistent headache and myalgia have been linked to stress during recovery [19]. Post-infectious fatigue is also common after dengue and can prolong functional impairment [20,22]. A recent comprehensive review of dengue infection further emphasised the importance of considering recovery and the broader consequences of dengue beyond its acute clinical manifestations [23]. Inflammatory and neuroimmune pathways have been proposed as additional mechanisms [6,7].

Lower anxiety among men could reflect sex-related differences in psychological response after dengue. Female sex has been associated with greater post-dengue stress and fatigue [19,20,22], and wider evidence also indicates sex and gender differences in anxiety prevalence [24]. Previous mental-health illness was associated with anxiety, consistent with wider NTD evidence showing that socioeconomic and social factors are important correlates of psychological morbidity [25,26]. Although our study combined widowed and divorced participants, previous systematic-review evidence has also shown increased mood and anxiety disorders following widowhood [27]. Unemployment or homemaking could reflect economic vulnerability, although these pathways were not directly measured.

Stress was associated with longer hospital stay, smoking, middle income, and rural residence. Longer admission could reflect greater disease burden, consistent with dengue-specific reviews describing disease severity and hospitalisation-related stress in relation to psychiatric sequelae [6,7]. The association between smoking and stress is also consistent with meta-analytic evidence showing an association between stress or emotional distress and smoking [28]. Socioeconomic disadvantage has been associated with mental-health problems across NTD populations [25,26]. However, evidence specifically linking middle income or rural residence with post-dengue stress remains limited, and these findings require confirmation. Previous COVID-19 infection was associated with stress, although evidence from Long COVID research [29] does not establish a dengue-specific mechanism. The association between August infection and anxiety also lacks an established explanation; both require investigation.

Strengths of this study include recruitment during hospitalisation followed by assessment at a predefined time approximately one month after discharge, allowing evaluation of an understudied stage of dengue recovery. The follow-up rate was high, with 288 of 306 eligible participants completing assessment (94.1%). Depression, anxiety, and stress were assessed simultaneously using the Bangla DASS-21, which has demonstrated satisfactory psychometric properties [13]. Use of the validated Bangla DASS-21 supported culturally and linguistically appropriate assessment [30]. The questionnaire was administered by trained research personnel, and completed assessments were reviewed by psychologists to enhance measurement consistency and scoring accuracy. The multivariable analyses incorporated sociodemographic, behavioural, and clinical factors. The regression analysis indicated no substantial multicollinearity, while the non-significant Hosmer–Lemeshow tests did not identify evidence of poor model calibration. Equal numbers of urban and rural participants also allowed residence to be examined in relation to psychological outcomes. This study adds evidence from Bangladesh on the co-occurrence of depressive, anxiety, and stress symptoms during early post-hospital recovery.

Several limitations should be considered. Psychological symptoms were not measured before dengue and no non-dengue comparison group was included, so the extent to which the observed symptoms were attributable specifically to dengue cannot be determined. Furthermore, psychological symptoms were assessed at a single time point approximately one month after discharge. As the DASS-21 captures symptoms experienced during the preceding seven days, the findings cannot establish when symptoms began, whether they persisted throughout recovery, or how they changed over time. DASS-21 measures symptom severity rather than providing psychiatric diagnoses; the findings therefore represent moderate- or-greater depressive, anxiety, and stress symptoms rather than diagnosed disorders [13]. Inflammatory and neuroimmune pathways have been proposed as possible mechanisms [6,7], but these were not assessed in this study. Smoking status, previous COVID-19 infection, and previous mental-health illness were self-reported, potentially introducing recall and reporting bias and increasing the risk of misclassification. Self-reported mental-health history may also have missed previously undiagnosed conditions. The single-centre tertiary-hospital setting, age restriction to 18–60 years, and exclusion of patients with chronic physical diseases limit generalisability and could introduce selection bias. Findings are most applicable to comparable hospitalised adults in Bangladesh and potentially similar dengue-endemic settings.

The high prevalence and co-occurrence of psychological symptoms in this study highlight the importance of considering mental health during post-dengue follow-up. Because 11.5% of participants experienced depression, anxiety, and stress concurrently, assessment across several psychological domains could identify broader distress than focusing on a single outcome. Positive screening results should be followed by clinical assessment rather than interpreted as psychiatric diagnoses. Future studies should use prospective multicentre designs with repeated assessment from the acute phase through recovery and should include comparison groups to better understand whether these psychological symptoms are specifically related to dengue. Measures of dengue severity, persistent symptoms, laboratory markers, previous psychiatric history, social support, economic consequences, and return to work should also be included. Longer follow-up is needed to distinguish short-term symptoms from persistent morbidity, particularly because some prospective evidence shows improvement over time [12], whereas population-based data indicate that some psychiatric outcomes remain elevated for longer periods [11].

In conclusion, depressive, anxiety, and stress symptoms were common approximately one month after dengue hospitalisation, often co-occurring. Associated factors differed across outcomes. These findings support attention to psychological well-being during recovery and further longitudinal research to identify persistent symptoms and higher-risk patients.

## Data Availability

The individual-level data underlying the findings of this study are not publicly available because they contain sensitive health and mental-health information collected from human participants. Public disclosure of these data could compromise participant confidentiality, particularly when sociodemographic and clinical variables are considered together. The study was approved by the Ethical Review Committee of the Diabetic Association of Bangladesh (BADAS-ERC/EC/24/08).

## Acknowledgments

We thank all participants who took part in the study. We also acknowledge the staff of BIRDEM General Hospital, Dhaka, Bangladesh, for their support with participant recruitment and follow-up, and the personnel associated with Thikana Psychiatric and Drug Addiction Clinic, Mohammadpur, Dhaka, for assistance with the psychological assessments.

